# Longitudinal Electrocochleography as an Objective Measure of Serial Behavioral Audiometry in Electro-Acoustic Stimulation Patients

**DOI:** 10.1101/2022.06.01.22275785

**Authors:** Viral D. Tejani, Jeong-Seo Kim, Christine P. Etler, Jeffrey Skidmore, Yi Yuan, Shuman He, Marlan R. Hansen, Bruce J. Gantz, Paul J. Abbas, Carolyn J. Brown

## Abstract

Minimally traumatic surgical techniques and advances in cochlear implant (CI) electrode array designs have allowed acoustic hearing present in a CI candidate prior to surgery to be preserved post-operatively. As a result, these patients benefit from combined electric-acoustic stimulation (EAS) post-operatively. However, 30-40% of EAS CI users experience a partial loss of hearing up to 30 dB after surgery. In the present study, electrocochleography (ECoG) was used to study cochlear microphonic (hair cell response) and auditory nerve neurophonic (neural response) in patients with preserved hearing and patients with loss of hearing. These measures were obtained longitudinally over the course of CI use. At each test session, ECoG amplitude growth functions for several low-frequency stimuli were obtained. The threshold, slope, and suprathreshold amplitude at a fixed stimulation level was obtained from each growth function at each time point. Subjects were categorized as having stable hearing or loss of hearing. Longitudinal linear mixed effects models were used study trends in ECoG thresholds, slopes, and amplitudes for these two categories of subjects. Results showed that CM and ANN thresholds and amplitudes were stable in CI users with preserved residual hearing. CM and ANN thresholds increased (worsened) while CM and ANN amplitudes decreased (worsened) for those with delayed hearing loss. The slope did not distinguish between subjects with stable hearing and subjects with delayed loss of hearing. These results provide a new application of post-operative ECoG as an objective tool to monitor residual hearing and understand the pathophysiology of delayed hearing loss.

## INTRODUCTION

Surgical techniques and cochlear implant (CI) electrode array designs have evolved in the last two decades to the extent that acoustic hearing present in a CI candidate prior to surgery can be preserved post-operatively. As a result, these patients benefit from combined electric-acoustic stimulation (EAS) post-operatively, where the electrode array provides electrical stimulation of the auditory nerve and an integrated hearing aid provides acoustic amplification. Since the proof of concept was demonstrated in the late 1990s (von Ilberg et al 1997; Gantz & Turner 2003), studies in the last two decades have consistently shown the benefits of EAS over conventional electrical-only stimulation for speech understanding in noise (Turner et al, 2004; Tejani & Brown 2020), perception of spectral and temporal cues (Gifford et al. 2008, 2010; Golub et al 2012; Tejani & Brown, 2020), music appreciation (Gfeller et al. 2006; Brockmeier et al. 2010), and localization (Dunn et al, 2010).

The benefits of EAS are maximized if residual acoustic hearing is successfully preserved long-term. It is critical for audiologists to conduct behavioral pure-tone audiometric testing longitudinally to monitor hearing thresholds and program the hearing aid portion of the EAS sound processors appropriately. Cochlear Nucleus L24 Hybrid CI user can generally maintain stable low-frequency audiometric thresholds (125, 250, and 500 Hz) of 50 - 70 dB HL up to five years post-surgery (Gantz et al, 2018; Roland et al, 2018) which is within the capabilities of the integrated hearing aid to provide amplification. However, about 30-40% of EAS CI users experience a partial loss up to 30 dB immediately and/or several months after surgery. In most cases, this additional hearing loss is not severe enough to preclude use of acoustic amplification (Lenarz et al. 2013; Van Abel et al. 2015; Scheperle et al. 2017; Pillsbury et al. 2018; Roland et al. 2018), but the loss of hearing can have a negative effect on performance and is an outcome that clinicians and researchers attempt to minimize.

Several theories are present as to why EAS CI users lose hearing post-operatively. Hearing loss immediately after surgery may result from insertion / structural trauma (Adunka et al, 2010). Delayed hearing loss that occurs months after surgery could be due to intracochlear fibrosis / osteoneogenesis (O’Leary et al. 2013; Quesnel et al. 2016; Foggia et al, 2019; Tejani et al, 2022) which could affect cochlear mechanics (Choi & Oghalai 2005). Histological data have also suggested compromised endocochlear potentials (Tanaka et al. 2014; Reiss et al. 2015) while not implicating hair cell / neural damage. Hair cell, neural, pre-synaptic ribbon counts, and post-synaptic receptor counts are stable post-loss of acoustic hearing (O’Leary et al. 2013; Tanaka et al. 2014; Reiss et al. 2015; Quesnel et al. 2016; but see Li et al. 2020 who reported cochlear neuropathy/synaptopathy).

To address some of these theories, our institution has used electrocochleography (ECoG) as an objective electrophysiological tool to assess peripheral auditory function in EAS CI users (Abbas et al. 2017; Kim et al. 2018; Tejani et al. 2019, 2021; see Eggermont 2017 for review on ECoG). The cochlear microphonic (CM) portion of the electrocochleogram reflects hair cell function, while the auditory nerve neurophonic (ANN) represents sustained phase-locked neural activity. Two other components of the electrocochleogram are the summating potential (SP) and the compound action potential (CAP). The SP presents as a baseline shift and may contain both hair cell and neural contributions (Pappa et al. 2019) while the CAP is a neural response occurring at the stimulus onset and offset. The underlying rational for use of ECoG in EAS studies is that both hair cell and neural function can be assessed in response to acoustic stimulation, thereby providing an objective measure of residual acoustic auditory function. Thus, changes in hair cell potentials and/or neural potentials with loss of residual hearing could shed light on hearing loss etiologies (e.g., Tejani et al, 2021).

Previous studies from our institution and others have validated ECoG as a potential tool to assess residual hearing and cochlear function. CM and ANN thresholds correlate strongly with behavioral audiometric thresholds (Abbas et al, 2017; Koka et al, 2017), which can aid with programming the hearing aid portion of the EAS sound processor (Agrawal et al, 2021). Intraoperative measures of ECoG are used to guide cochlear implantation to minimize cochlear trauma (Bester et al, 2022; Lenarz et al, 2022) and have been shown to correlate with post-operative speech understanding (Fontenot et al, 2019; Canfarotta et al, 2021; Walia et al, 2022). One important aspect of ECoG that hasn’t been sufficiently validated is the long-term stability of these measures. Our previous studies have mainly focused on test-retest reliability of ECoG thresholds at two time points for EAS patients with stable hearing and EAS patients with loss of hearing. These studies also presented limited longitudinal suprathreshold ECoG amplitude data (Abbas et al, 2017; Kim et al 2018; Tejani et al, 2019). These data did indeed show that ECoG measures are stable at two time points for EAS patients with stable hearing while changes in ECoG did mirror changes in behavioral audiometry for EAS patients with fluctuating acoustic hearing or loss of acoustic hearing. However, if ECoG is to be used as an objective method of monitoring residual hearing over the course of EAS CI use, then repeated measures of these potentials should remain stable over time for EAS CI users with stable post-operative hearing preservation. Additionally, changes in behavioral audiometry for EAS CI users with loss of residual hearing should also be reflected in changes in ECoG measures.

## METHODS

This study was approved by the University of Iowa Institutional Review Board (IRB 201805740). Subjects signed an informed consent form.

### Subject Population and Classification

A pool of 40 subjects implanted with Cochlear Corporation electrode arrays were included in this study. They were all implanted between the years 2006 and 2021. All subjects were adults who presented with significant residual acoustic hearing at time of implantation. The average pre-operative low-frequency pure tone average (PTA) of 125 to 500 Hz was 40.88 ± 13.12 dB HL.

Subjects were seen at several time points post-operatively for both behavioral audiometry and ECoG recordings. Time points included 0.5, 1, 3, 6, 12 months, and annually after 12 months, though not all subjects were tested at all time points and there were subjects enrolled into the study years post-surgery. While behavioral audiometry was done at initial activation, ECoG recordings were not done at that time point. Changes in pure-tone audiometric thresholds relative to the baseline appointment were used to classify subjects into two groups: one group of subjects with stable acoustic hearing and another group of subjects with loss of acoustic hearing. The baseline appointment was the first appointment at which both audiometry and ECoG recordings were done. For purposes of subject classification, loss of acoustic hearing was defined as a > 5 dB decline in behavioral threshold between the subject’s most recent appointment and the baseline appointment. We focused on 250, 500, 750, and 1000 Hz audiometry and, as explained in more details in a subsequent section, we conducted analyses for each frequency separately. Thus, the number of subjects in the stable vs hearing loss group may differ for each frequency.

Table 1 describes subject demographics in more detail, including array type, classification into stable vs hearing loss group, the use of intraoperative ECoG monitoring, and electrode insertion depth. The last two variables are explained in the next section.

**Table 1:** Subject Demographics.

| Subject ID | Ear | Pre-op PTA<br>(125 - 500 Hz) | Array | Implant<br>Date | ECoG<br>monitoring? | Full or<br>Partial Insertion? | Time points tested<br>post activation<br>(months) | 250 Hz<br>Category | 500 Hz<br>Category | 750 Hz<br>Category | 1000 Hz<br>Category |
| --- | --- | --- | --- | --- | --- | --- | --- | --- | --- | --- | --- |
| A8 | R | 18 | S8 | 05/2006 | N | Full | 156, 169 | S | S | S | S |
| L3R | R | 45 | L24 | 5/2010 | N | Full | 109, 122 | HL | HL | S | S |
| T10L | L | 27 | S12 | 12/2010 | N | Full | 100, 113 | S | S | S | S |
| L13L | L | 52 | L24 | 11/2012 | N | Full | 72,81,85 | S | S | S | S |
| T12L | L | 20 | S12 | 11/2013 | N | Full | 54, 72 | S | HL | S | S |
| L17L | L | 37 | L24 | 11/2013 | N | Full | 72, 84 | S | S | S | S |
| L25L | L | 42 | L24 | 8/2014 | N | Full | 55, 60 | S | S | S | S |
| S35L | L | 48 | 422 | 9/2014 | N | Full | 60, 72 | S | S | S | S |
| S44R | R | 58 | 422 | 2/2015 | N | Full | 37, 49 | S | S | HL | S |
| S36R | R | 55 | 422 | 2/2015 | N | Full | 36, 48 | S | S | S | S |
| L33L | L | 25 | L24 | 5/2015 | N | Full | 49, 61 | S | S | S | S |
| L44R | R | 30 | L24 | 12/2015 | N | Full | 36, 48 | S | HL | HL | S |
| L43R | R | 43 | L24 | 6/2016 | N | Full | 36, 48 | S | S | HL | S |
| S12RW-3L | L | 21 | S12RW | 12/2016 | N | Full | 6, 12 | S | S | S | S |
| L66R | R | 40 | L24 | 4/2017 | N | Full | 12, 22 | HL | S | S | S |
| L67R | R | 47 | L24 | 5/2017 | N | Full | 23, 38 | S | S | S | S |
| L77R | R | 35 | L24 | 8/2017 | N | Full | 26, 35 | HL | HL | HL | HL |
| L80R | R | 22 | L24 | 5/2018 | Y | Full | 0.5, 1, 3 | S | S | S | S |
| L81R | R | 27 | L24 | 6/2018 | Y | Full | 0.5, 1, 3, 6 | HL | HL | HL | HL |
| S12RW-6R | R | 33 | S12RW | 10/2018 | Y | Full | 0.5, 1, 3, 6, 12 | S | HL | HL | HL |
| L86R | R | 41 | L24 | 1/2019 | Y | Full | 0.5, 1, 3, 6 | S | S | S | S |
| 522-11L | L | 37 | CI522 | 7/2019 | Y | Full | 0.5, 1 | S | S | S | S |
| 622-2R | R | 63 | CI622 | 11/2019 | Y | Full | 0.5, 1, 3 | S | S | S | S |
| 622-3R | R | 47 | CI622 | 11/2019 | Y | Full | 0.5, 1, 3 | S | S | S | S |
| 624-20R | R | 35 | CI624 | 1/2020 | Y | Full | 0.5, 1, 3, 6, 12 | HL | HL | HL | HL |
| 622-5R | R | 43 | CI622 | 1/2020 | Y | 3 Electrodes<br>Extracochlear | 0.5, 1, 6, 12 | S | S | S | S |
| 624-1R | R | 46 | CI624 | 5/2020 | Y | Full | 0.5, 1, 3, 9 | HL | HL | HL | HL |
| 624-2R | R | 60 | CI624 | 5/2020 | Y | Full | 0.5, 1, 3, 6 | HL | HL | HL | HL |
| 624-4R | R | 65 | CI624 | 6/2020 | Y | Full | 0.5, 1, 3, 8 | HL | HL | HL | HL |
| 624-6R | R | 53 | CI624 | 6/2020 | Y | Full | 0.5, 1, 3, 6 | S | S | S | S |
| 624-8R | R | 43 | CI624 | 7/2020 | Y | Full | 0.5, 1, 7 | S | S | S | S |
| 624-9L | L | 55 | CI624 | 7/2020 | Y | Full | 0.5, 1, 3, 6 | S | S | S | S |
| 624-11R | R | 43 | CI624 | 8/2020 | Y | Full | 0.5, 1 | S | S | S | S |
| 624-12L | L | 35 | CI624 | 09/20 | Y | Full | 0.5, 1, 3, 12 | HL | HL | S | S |
| 624-13L | L | 30 | CI624 | 10/2020 | Y | Full | 0.5, 1, 3, 8 | HL | HL | HL | HL |
| 624-15R | R | 52 | CI624 | 11/2020 | Y | Full | 0.5, 1 | S | S | S | S |
| 624-16L | L | 62 | CI624 | 12/2020 | Y | Full | 0.5, 1, 3 | HL | HL | S | S |
| 624-18L | L | 27 | CI624 | 12/2020 | Y | Full | 0.5, 1, 3 | S | HL | S | S |
| 624-19R | R | 20 | CI624 | 12/2020 | Y | 3 Electrodes<br>Extracochlear | 0.5, 1 | S | S | HL | S |
| 624-21L | L | 53 | CI624 | 3/2021 | Y | Full | 0.5, 1, 3, 6 | S | S | S | S |

### Cochlear Implantation

Various Cochlear Corporation electrode arrays were used in this study, including the S8 / S12 / L24 Hybrids arrays, CI 422/522/622 slim lateral wall arrays, and the CI 624 Slim 20. Table 2 describes the arrays in detail, including length, insertion depth, and number of electrodes.

**Table 2:**
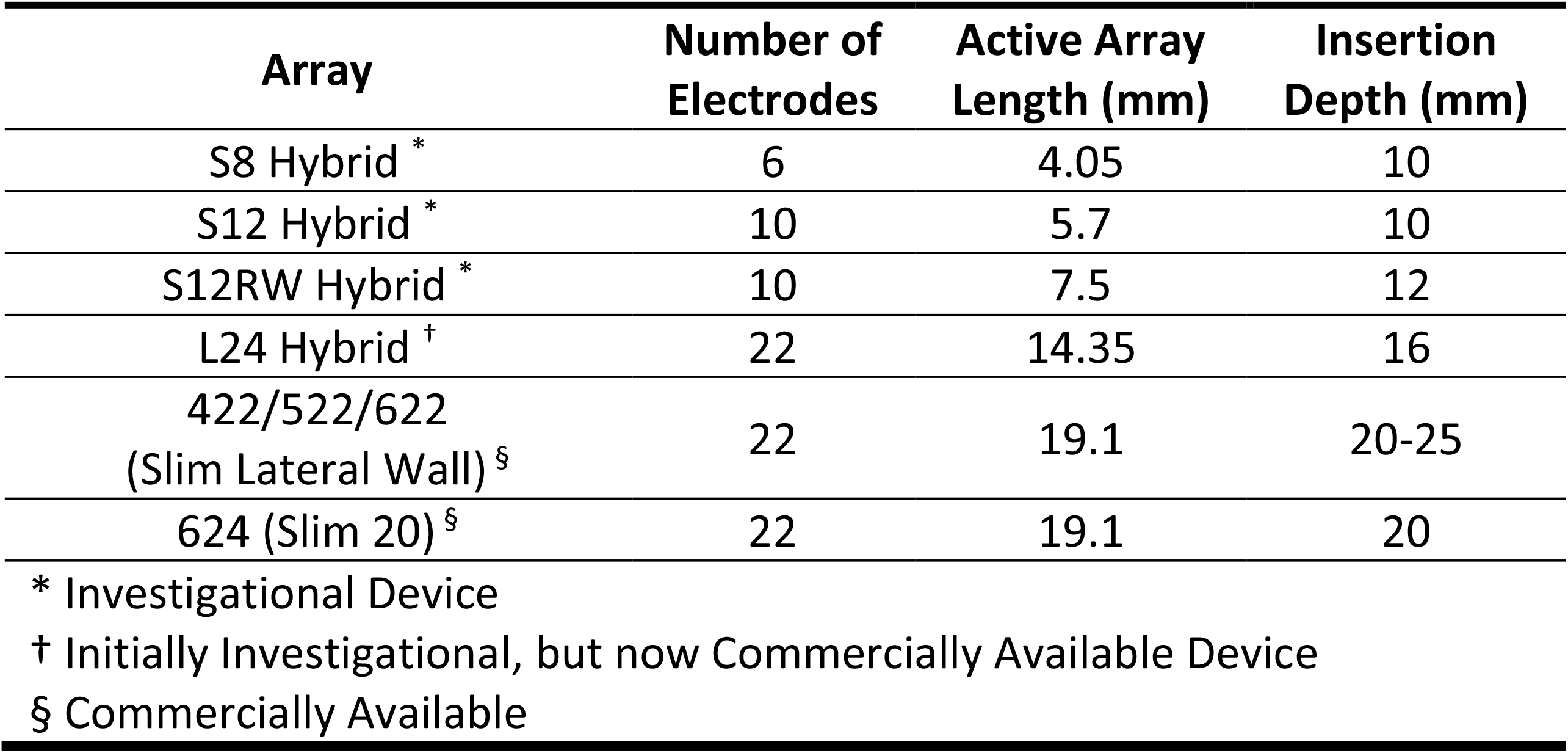
Hybrid and Standard Electrode Array Dimensions.

Implantation was performed by authors BJG and MRH at the University of Iowa Hospitals and Clinics under hearing preservation protocols using soft insertion techniques. Subjects received intravenous dexamethasone during surgery, a 1-week course of prednisone (1 mg/kg/day) beginning immediately postoperatively, and a second 1-week course beginning the day prior to activation of the CI. The steroids were an attempt to reduce inflammatory responses and subsequent loss of hearing that may be associated with surgical trauma (e.g., Rauch et al. 2011). Subjects implanted prior to 2013 were implanted via a cochleostomy while later subjects were implanted via the round window in attempts to minimize cochlear trauma (e.g., Adunka et al. 2004). At the end of insertion, the electrode array was secured to the tegmen mastoideum to reduce chances of electrode migration and ensure stability of the intracochlear array. Cochlear implantation of recent patients was also done in conjunction with intraoperative ECoG measures to guide insertion trajectory and depth (Lenarz et al, 2021; Bester et al, 2022); in two cases partial electrode insertions were purposely performed to attempt to preserve residual acoustic hearing. Table 1 indicates which subjects were implanted in conjunction with intraoperative ECoG and which two subjects had partial insertions performed.

### Electrococheography Recordings

Recordings were performed using a custom in-house system that utilizes Python programming and version 2 of the Nucleus Implant Communicator (NIC) routines. These programs were used to trigger acoustic stimulation and record a response from an intracochlear electrode (see Tejani et al, 2019 for specific details on software and hardware adaptations). The stimuli were low-frequency tone bursts that were presented to the implanted ear via an insert earphone. Presentation levels ranged from below behavioral detection threshold to the maximal comfort level. A response was recorded from the most apical electrode in the array. Stimuli were 24-ms tone bursts with a 1-cycle rise / fall time, or 1 ms, whichever was longer, shaped by a cosine-squared window. Stimuli were presented at a 10-Hz stimulation rate in both condensation and rarefaction polarities. Stimuli frequencies were 250, 500, 750, and 1000 Hz.

Recordings were repeated at several time points after activation of the CI, typically coinciding with clinical checkup appointments. Time points included 0.5, 1, 3, 6, 12 months, and annually after 12 months. Exact time points varied based on each subject’s availability. Subjects were also seen for interim appointments if they experienced a loss of acoustic hearing. Additionally, while recordings were attempted for all stimulus frequencies, we focused on 500 Hz if there were time / subject availability limitations.

### Statistical Analysis

ECoG recordings were done at several levels from behavioral threshold level to the maximal comfort levels. CM and ANN amplitudes were plotted as a function of stimulus level to construct amplitude growth functions. As detailed in the “Electrococheography Amplitude Growth Functions” section of the results section, three parameters were extracted – threshold, slope, and suprathreshold amplitude at a fixed level. These parameters were extracted for all amplitude growth functions at all test frequencies that were collected at every appointment. Changes in threshold, slope, and amplitude at each appointment were calculated relative to baseline. These three metrics served as the dependent variable as part of a linear mixed effects model (LME). The change in each dependent variable over time was evaluated separately with LME models for two groups of subjects: subjects with stable hearing and subjects with loss of hearing. For each LME model the deviation from baseline of the dependent variable was the response variable, the fixed effect was time from baseline, and subject was the random effect. No intercepts were included in the model to force the model to pass through the baseline datapoint. In other words, only slope was considered for the fixed and random effects. Significance values were adjusted using the False Discovery Rate to minimize potential Type I errors from repeated analyses.

## RESULTS

### Electrococheography Amplitude Growth Functions

Responses to condensation and rarefaction stimuli were subtracted from one another to emphasize the CM and added to one another to emphasize the ANN (Aran and Charlet de Sauvage 1976; Henry 1995; Lichtenhan et al. 2013). Since this difference and summation technique does not result in a pure separation of CM and ANN (Forgues et al. 2014; Abbas et al. 2017), we hereafter refer to the potentials as CM/DIFF and ANN/SUM. A Fast Fourier Transform (FFT) analysis of the time domain data into the frequency domain was then performed, resulting in a resolution of 37.74 Hz/bin.

Figure 1 shows an example recording from one subject (622-5R) who was seen at his 12-month appointment. The top left panel shows the resulting response to both condensation and rarefaction polarities for a 500 Hz tone burst. The top middle and top right panels show the resulting CM/DIFF and ANN/SUM recordings. The CM/DIFF waveform oscillates at a 500 Hz frequency while the ANN/SUM waveform oscillates at a 1000 Hz frequency. The doubling in frequency present in ANN/SUM recordings results from activation of nerve fibers responding to the depolarizing phase of the stimulus. There is a half-cycle difference in latency of the depolarizing phase for rarefaction and condensation stimuli; when both recordings are summed, the resulting waveform oscillates at twice the stimulus frequency. In this case, there is also evidence of a compound action potential in the ANN/SUM recording, with a latency of about 4 ms. The bottom middle and bottom right panels show the resulting FFT for each recording. Note there is a peak at the stimulus frequency for the CM/DIFF and a peak at twice the stimulus frequency for the ANN/SUM. In addition, higher order harmonics are sometimes present due to distortions in the hair cell and neural signal transduction process (Forgues et al, 2014).

**Figure 1:**
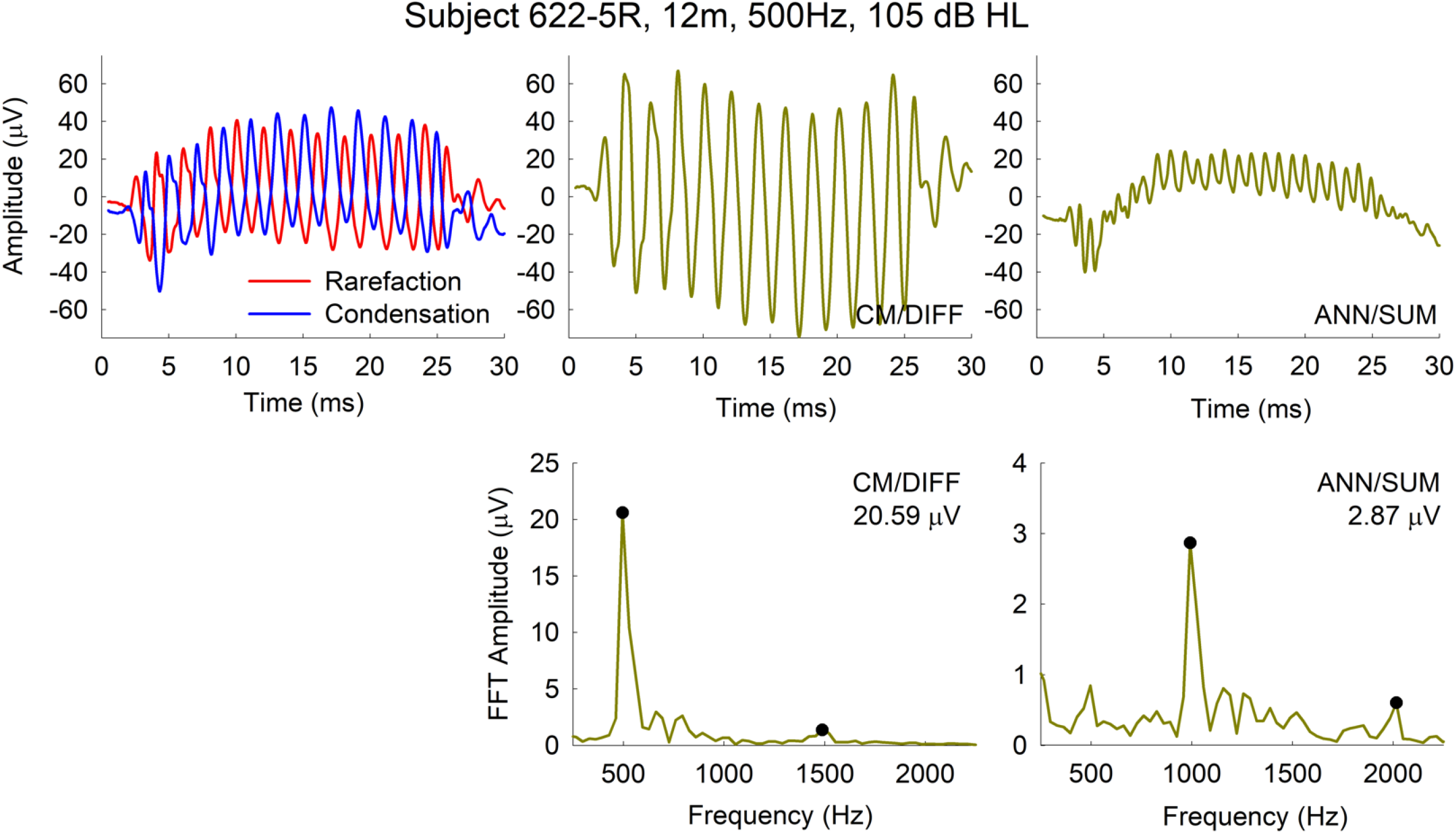
Example ECoG recording for subject 622-5R in response to a 500 Hz tone burst presented at 105 dB HL. The 12m indicates he was seen at his 12-month post CI appointment. Top left panel shows the responses to rarefaction and condensation stimuli. Top middle and right panels show the resulting CM/DIFF and ANN/SUM recordings. The bottom panels represent the resultant FFT analyses of the CM/DIFF and ANN/SUM recordings. Significant FFT peaks (including higher order harmonics) are marked by the black filled circles, with the corresponding FFT amplitudes indicated in the top right of the FFT plots.

Figure 2 shows amplitude growth functions for the CM/DIFF and ANN/SUM recordings for the same subject (622-5R). The peaks of the FFT at the stimulus frequency for the CM/DIF recordings were used in plotting the amplitude growth functions. This was similarly done for the peak at twice the stimulus frequency for the ANN/SUM growth functions. As previously mentioned, we extracted threshold, slope, and a suprathreshold amplitude. Note that this results in 12 variables extracted for the CM and 12 variables extracted for the ANN (4 frequencies x 3 variables), all of which were subjected to individual LME analyses. The threshold is the lowest stimulus level that results in an ECoG response, measured in dB HL. The slope represents the rate of change in amplitude as the stimulus level increases, calculated using a linear regression, and measured in μV/dB. For the suprathreshold amplitude, we identified the highest stimulus level that was used across all time points for a particular stimulus frequency. We then extracted the corresponding electrocochleogram amplitudes for all those time points. For example, in the case of subject 622-5R represented in Figure 2, the highest level used for the 250 Hz stimulation frequency across all time points was 90 dB HL. The corresponding amplitude in the growth function for that stimulus level was used in the longitudinal analysis. Similarly, amplitudes corresponding to a 100, 110-, and 110-dB HL stimulus levels for 500, 750, and 1000 Hz stimulation frequencies were used in the analysis for this subject.

**Figure 2:**
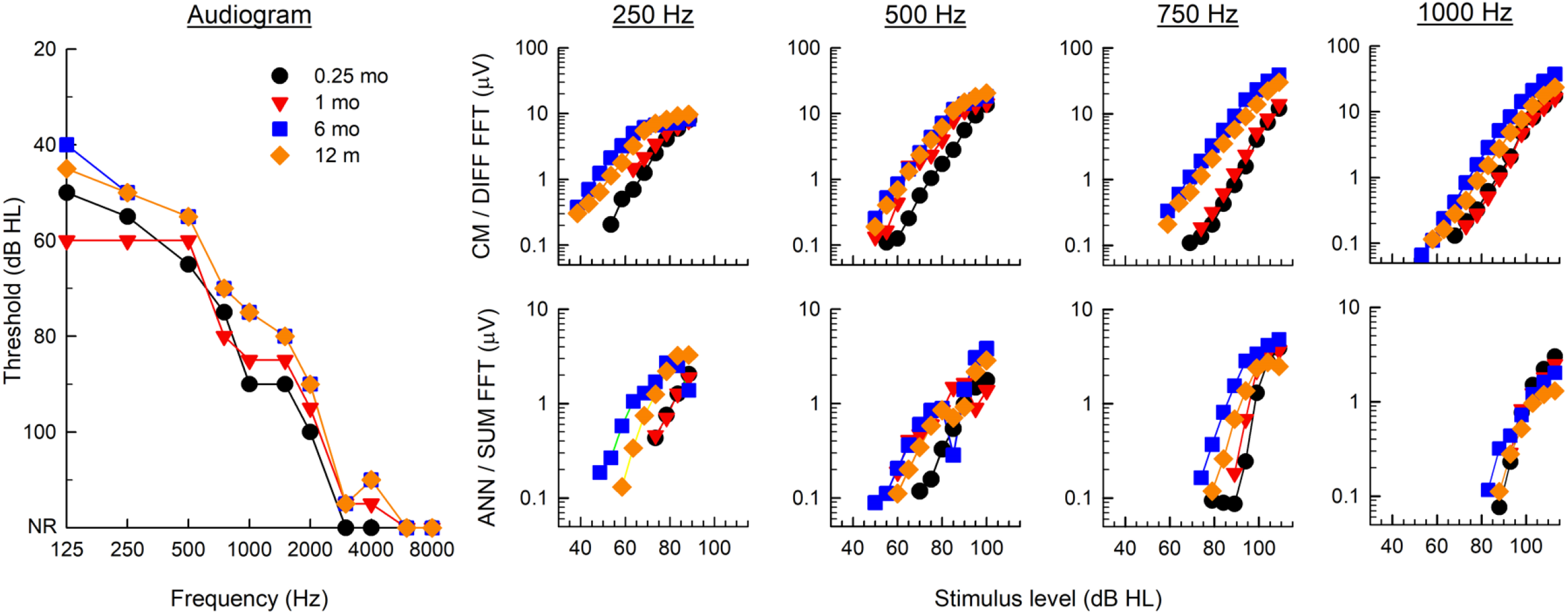
Longitudinal Audiograms and ECoG amplitude growth functions for Subject 622-5R. The left panel shows the behavioral audiograms. The top row of mini-panels show CM/DIFF amplitude growth functions while the bottom row of mini-panels show ANN/SUM amplitude growth functions for the four stimulus frequencies.

Figure 3 shows example longitudinal thresholds, slopes and amplitudes for the same subject represented in Figure 2. The top panels show raw values while the bottom panels show changes in values over time. In this case, the subject showed some improvement in audiometric hearing over time; this is likely due to resolution of middle ear fluid and resulting conductive hearing loss, as commonly seen immediately post CI surgery. As more clearly seen in the bottom panels, the changes in ECoG thresholds and amplitudes mirror changes in behavioral audiometric thresholds. Thresholds and amplitudes are generally stable during periods of stable hearing. Thresholds and amplitudes worsen with loss of hearing, and vice-versa.

**Figure 3:**
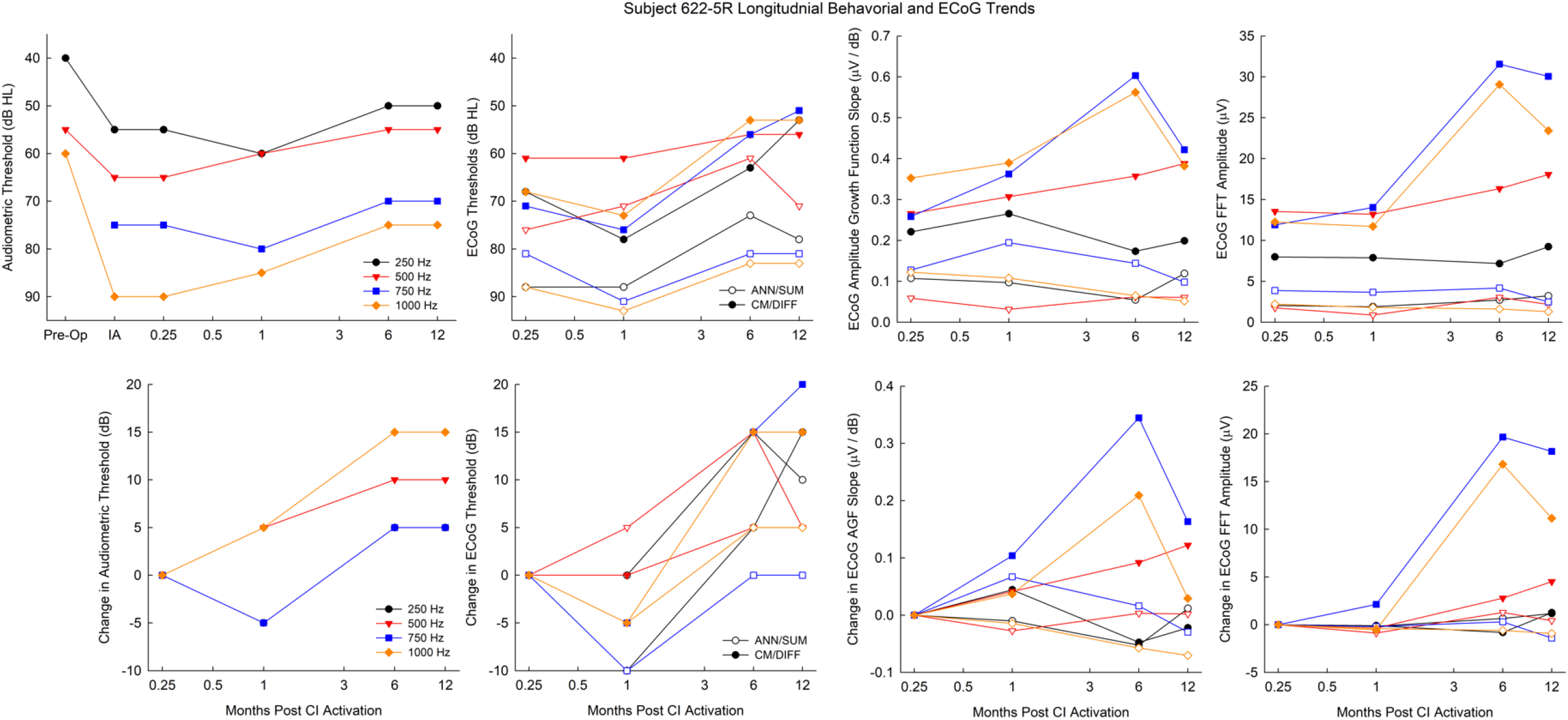
Longitudinal audiometric thresholds, ECoG thresholds, slopes, and amplitudes for subject 622-5R. The top row shows the raw values while the bottom row shows changes in these metrics relative to the baseline appointment conducted at 0.25 months post CI activation. For the ECoG metrics, closed symbols represent the CM/DIFF recordings while the open symbols represent the ANN/SUM recordings.

### Stability of ECoG measures

As previously mentioned, the primary question of interest was evaluating the stability of ECoG metrics over time in the stable hearing group and the hearing loss group. Thus, the change in each ECoG metric was calculated relative to baseline for each subject (e.g bottom panels of Figure 3). These metrics were obtained for all subjects, and separate LME models for each ECoG metric at each frequency were conducted to analyze whether longitudinal trends were statistically significant.

Table 3 summarizes the results of the LME models. For those with stable hearing, there are no changes noted in CM/DIFF or ANN/SUM thresholds or amplitude over time for any frequencies (except for the 250 Hz CM/DIFF threshold). In contrast, there are changes in thresholds and amplitudes noted in most cases for those with loss of hearing. As evidence by the *β* -values, amplitudes generally decreased (*β* < 0) and thresholds generally increased (*β* > 0) with loss of hearing. Additionally, though there was some evidence of the slope becoming shallower for the hearing loss group, (*β* < 0), the slope metric was generally not statistically significant.

**Table 3:**
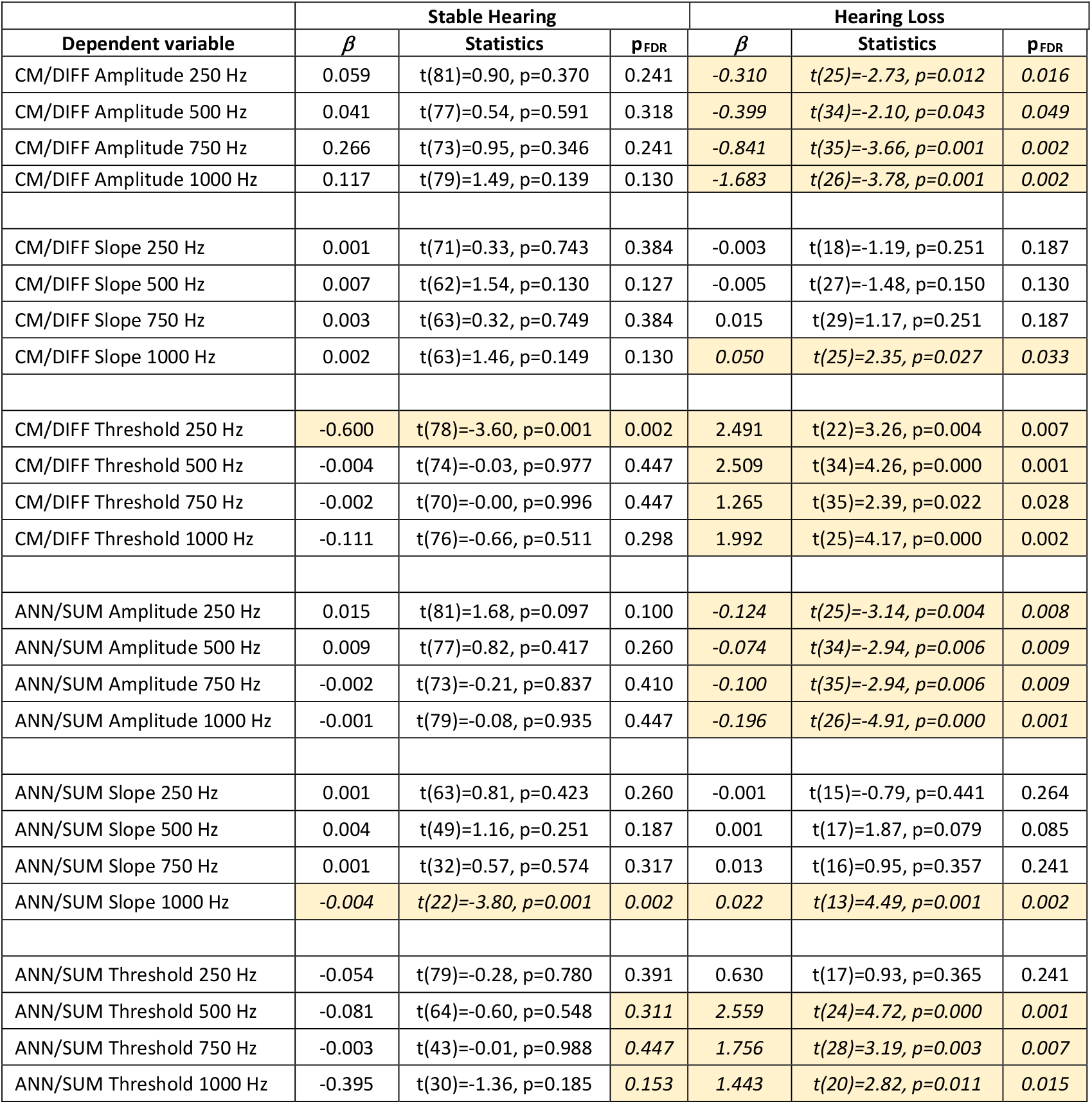
Results of LME analysis. P-values were adjusted using a False Discovery Rate. Italicized numbers indicate results that are statistically significant.

### Comparisons of CM/DIFF and ANN/SUM losses

It is evident from the previous analysis that both CM/DIFF and ANN/SUM metrics decline with declines in audiometric hearing. It was also of interest to see if there are equal drops in both CM/DIFF and ANN/SUM potentials, or if there is a greater decline in one potential compared to the other. If there is a greater decline in one potential, it may shed more insight into the physiology of delayed hearing loss.

Figure 4 plots changes in the CM/DIFF metric relative to the changes in ANN/SUM metric that occurs at the time point where hearing loss was identified. The two metrics of interest were threshold and amplitude, as those metrics were affected by loss of hearing. The slope of the ECoG amplitude growth function was not analyzed here since it was not sensitive to loss of acoustic hearing.

**Figure 4:**
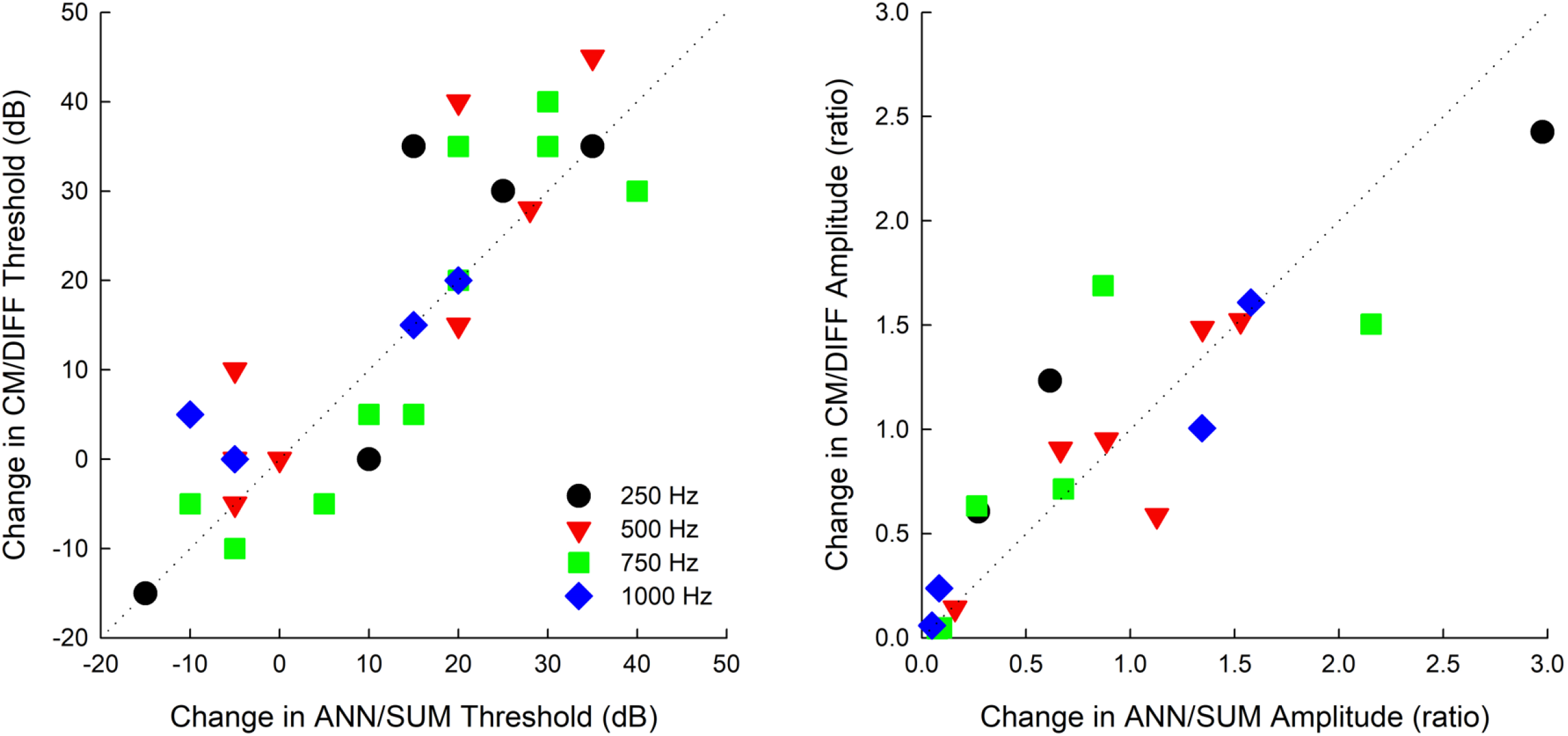
Comparisons of CM/DIFF and ANN/SUM changes after loss of hearing. The dotted line indicates equal changes in threshold (left panel) and amplitude (right panel). For the left panel, positive numbers indicate an increased (worsened) threshold with loss of hearing. For the right panel, a ratio < 1 indicates a decreased (worsened) amplitude with loss of hearing.

For the threshold analysis, the difference in the CM/DIFF threshold at the time point of hearing loss and the previous appt was calculated. The same calculation was performed for the ANN/SUM threshold.

These two differences were then plotted against one another. For the amplitude analysis, a ratio of the CM/DIFF amplitude at loss of hearing relative to the previous appointment was calculated. The same ratio calculation was performed for the ANN/SUM amplitudes. These two ratios were plotted against one another.

Figure 4 left panel focuses on threshold changes. As summarized in Table 4, there is a significant correlation between CM/DIFF threshold changes and ANN/SUM threshold changes. More importantly, a paired t-test compared the CM/DIFF and ANN/SUM threshold change at time point of hearing loss and showed that there were no differences in threshold changes. A similar result was found for the amplitude changes, in that both CM/DIFF and ANN/SUM amplitudes have similar decrements after loss of hearing.

**Table 4:** Comparisons of CM/DIFF and ANN/SUM changes after loss of hearing. Italicized numbers indicate results that are statistically significant

|  | Threshold Changes |  |  |  |  |  | Amplitude Changes |  |  |  |  |  |
| --- | --- | --- | --- | --- | --- | --- | --- | --- | --- | --- | --- | --- |
|  | Correlation |  |  | t-test |  |  | Correlation |  |  | t-test |  |  |
| Frequency (Hz) | *n* | *r* | *p* | *t* | *df* | *p* | *n* | *r* | *p* | *t* | *df* | *P* |
| 250 | 5 | 0.883 | *0.024* | -0.612 | 4 | 0.573 | 3 | 0.974 | *0.073* | -0.384 | 2 | 0.738 |
| 500 | 8 | 0.892 | *0.001* | -1.843 | 7 | 0.108 | 6 | 0.861 | *0.014* | 0.183 | 5 | 0.862 |
| 750 | 10 | 0.886 | *0.000* | 0.176 | 9 | 0.864 | 6 | 0.79 | *0.031* | -0.41 | 5 | 0.699 |
| 1000 | 4 | 0.93 | *0.035* | -1.414 | 3 | 0.252 | 4 | 0.969 | *0.015* | 0.338 | 3 | 0.758 |

### Correlations between Behavioral Audiogram, ECoG Slopes, ECoG Thresholds, and ECoG amplitudes

As a secondary analysis, CM/DIFF, ANN/SUM, and behavioral audiometric thresholds from each subject’s latest test session were obtained. Figure 5 shows correlations between behavioral audiometric thresholds and electrophysiologic thresholds. In general, electrophysiological thresholds are well correlated with behavioral thresholds. In addition, 500 and 750 Hz CM/DIFF thresholds are closest to behavioral thresholds. Note that for these correlational analyses, cases of no ECoG responses (as indicated by the open symbols) were plotted as 120 dB HL but were excluded from correlational analyses.

**Figure 5:**
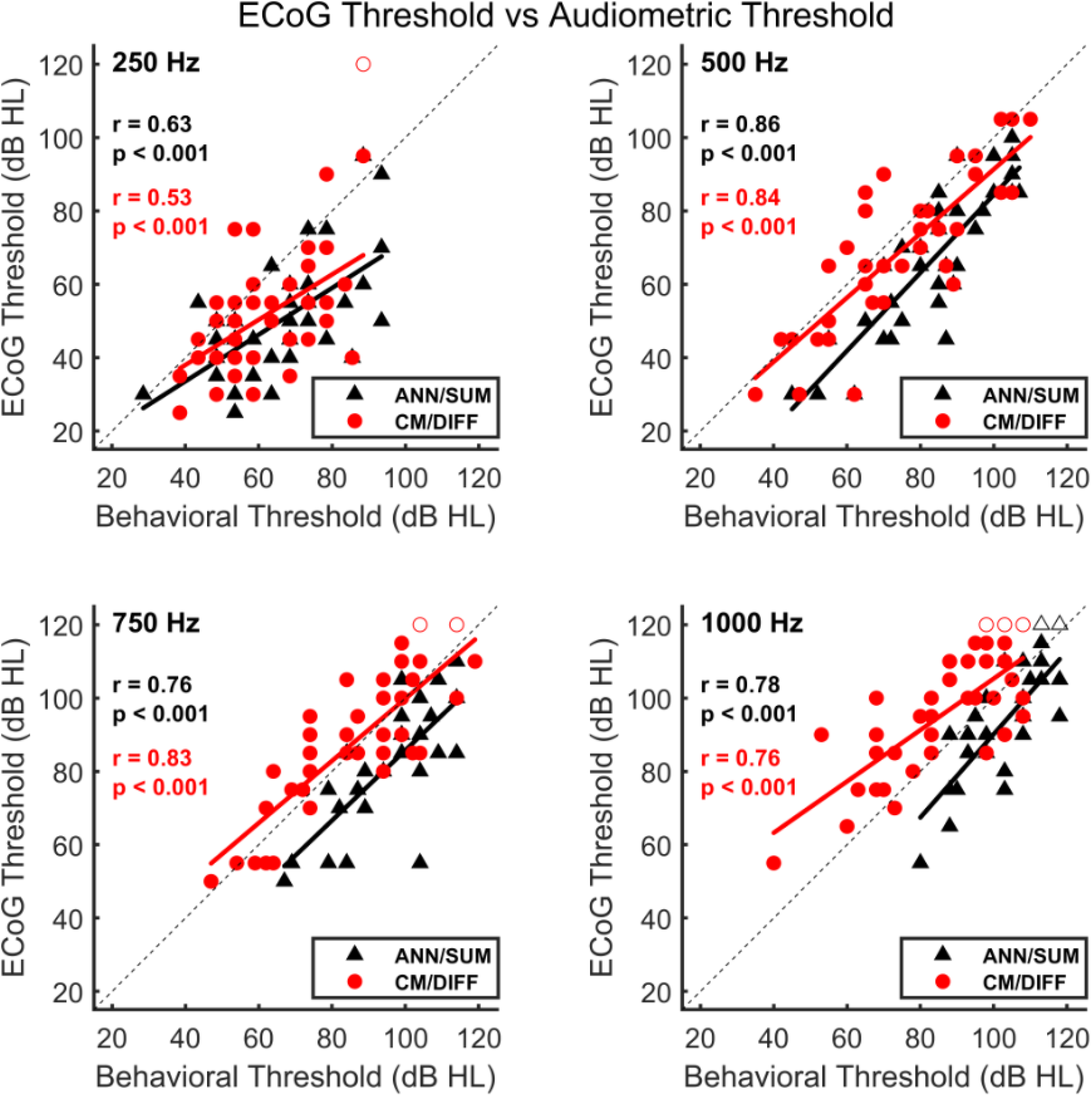
Correlations between ECoG and behavioral thresholds for 250, 500, 750, and 1000 Hz stimuli. The dotted line indicates equal CM/DIFF and ANN/SUM thresholds. Note that cases of no ECoG responses are indicated by the open symbols and are not included in correlational analyses.

Figure 6 shows correlations between behavioral audiometric thresholds and electrophysiologic slopes of the ECoG amplitude growth functions. It was thought that those with better residual hearing would have steeper slopes. In general, this was not the case, except for the 750 and 1000 Hz CM/DIFF growth slopes and the 1000 Hz ANN/SUM growth slopes (*r* = -0.407, -0.6654, and -0.5302, respectively, with *p* < 0.01 in all three case). Note that for these correlational analyses, cases of no behavioral audiometric responses (as indicated by the open symbols) were plotted as 120 dB HL but were excluded from correlational analyses.

**Figure 6:**
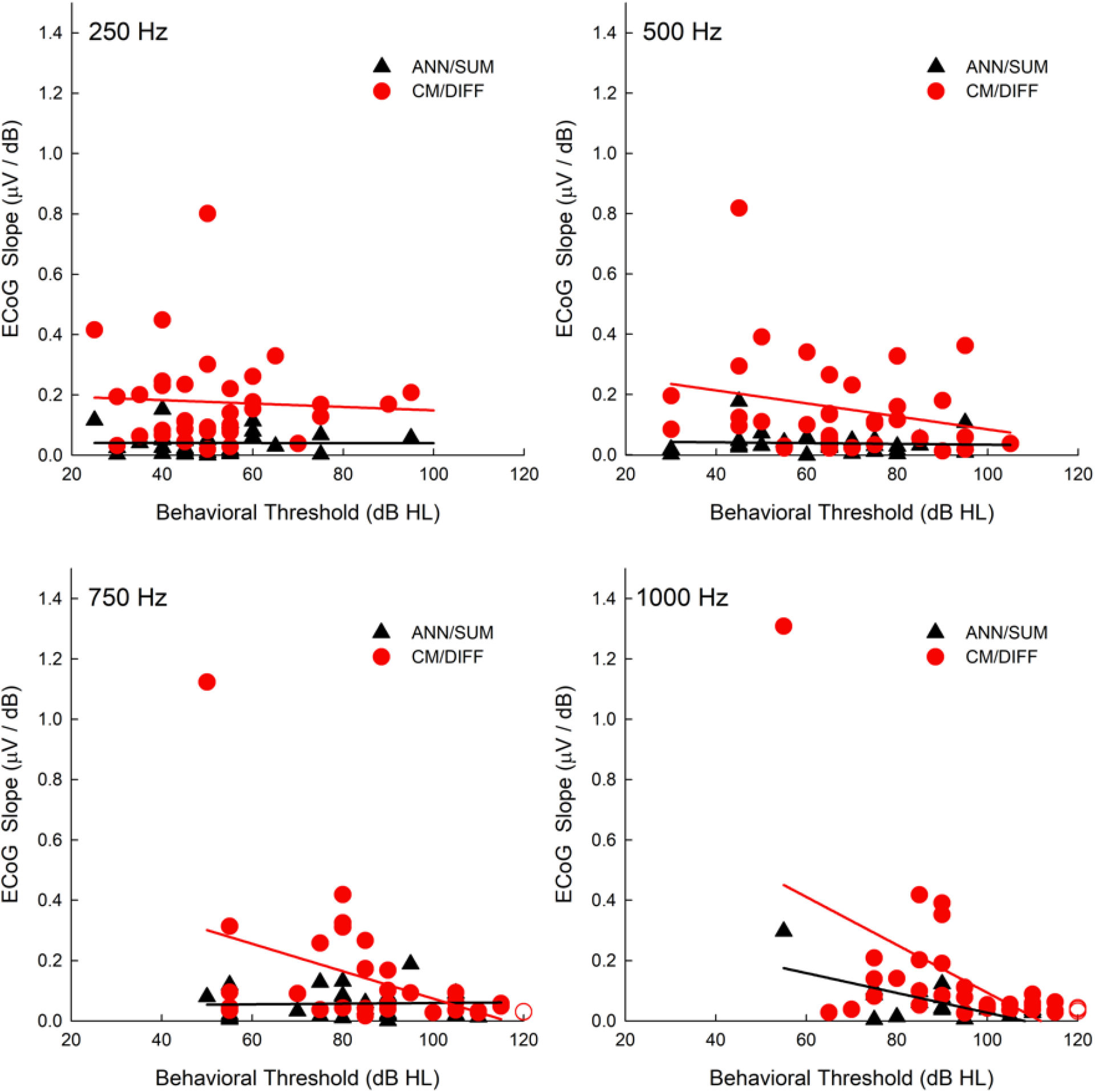
Correlations between ECoG slope and behavioral thresholds for 250, 500, 750, and 1000 Hz stimuli. Note that in the bottom plots, cases of no behavioral responses are indicated by the open symbols and are not included in correlational analyses.

Figure 7 shows correlations between behavioral audiometric thresholds and electrophysiologic amplitudes obtained using the subject’s C-level. It was thought that those with better residual hearing would have higher ECoG amplitudes. Generally speaking, there were trends of higher ECoG amplitudes for those with better residual hearing (lower audiometric thresholds). As summarized in Table 5, these trends were statistically significant or borderline statistically significant for the CM/DIFF potentials at all tested frequencies. These trends were statistically significant for the 500 Hz and 1000 Hz ANN/SUM potentials.

**Figure 7:**
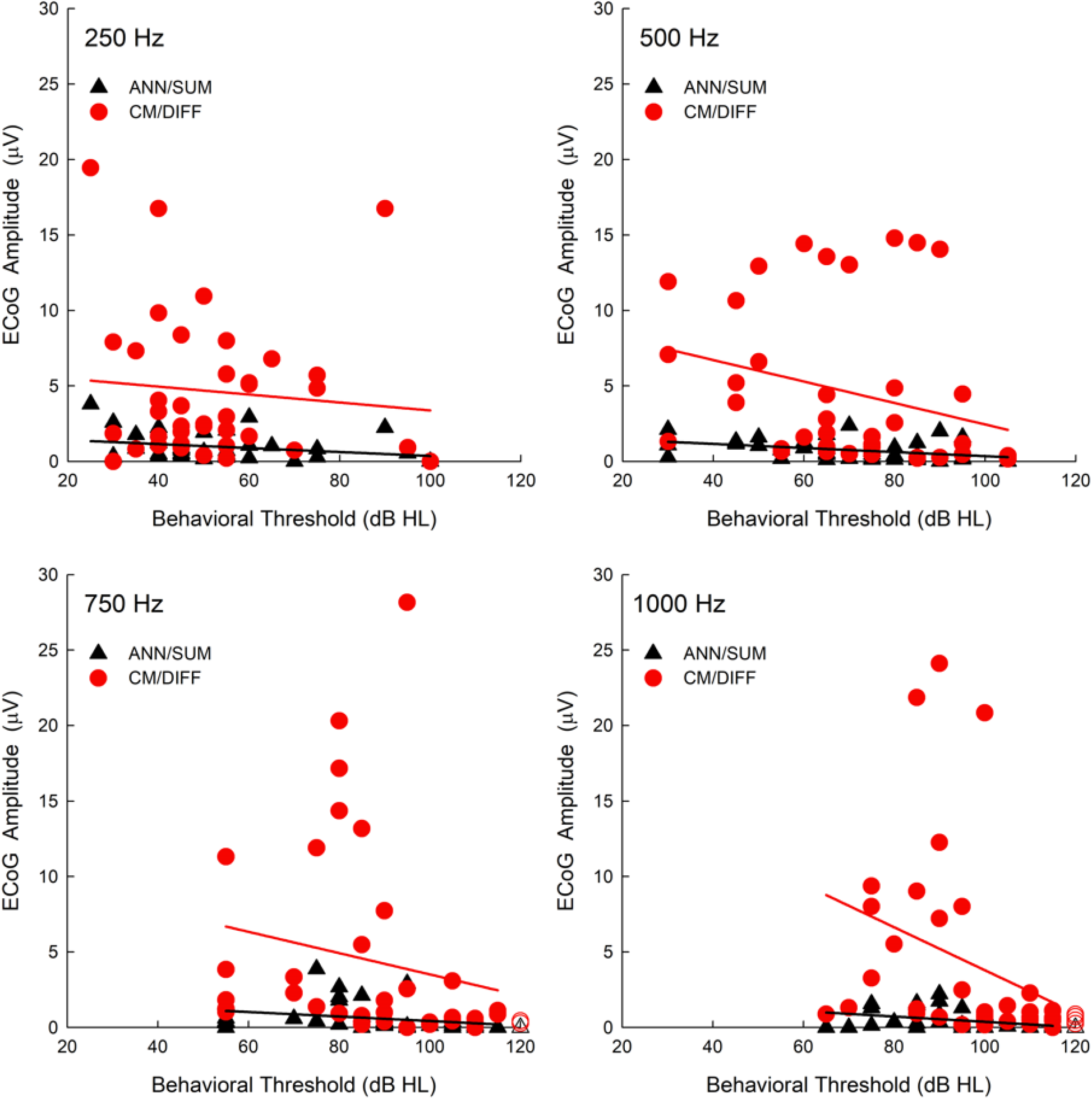
Correlations between ECoG amplitudes and behavioral thresholds for 250, 500, 750, and 1000 Hz stimuli. Note that in the bottom plots, cases of no behavioral responses are indicated by the open symbols and are not included in correlational analyses. Subject S12RW-6R was excluded from the correlational analysis as he was a clear outlier with unusually large (100 μV) amplitudes.

**Table 5:** Correlations between behavioral audiograms and suprathreshold ECoG amplitudes

| Frequency (Hz) | Correlation |  |  |
| --- | --- | --- | --- |
|  | *n* | *r* | *p* |
| 250 Hz CM/DIFF | 39 | 0.256 | 0.061 |
| 500 Hz CM/DIFF | 40 | 0.419 | *0.004* |
| 750 Hz CM/DIFF | 37 | 0.270 | 0.055 |
| 1000 Hz CM/DIFF | 34 | 0.391 | *0.011* |
| 250 Hz ANN/SUM | 39 | 0.095 | 0.285 |
| 500 Hz ANN/SUM | 39 | 0.280 | *0.042* |
| 750 Hz ANN/SUM | 37 | 0.185 | 0.140 |
| 1000 Hz ANN/SUM | 35 | 0.111 | *0.035* |

The slopes of the CM/DIFF and ANN/SUM amplitude growth functions were plotted against one another to ascertain if there were correlations between both and to understand if one potential grows faster than the other. From Figure 8, it does appear that both metrics are correlated. More importantly is that the CM/DIFF grows more quickly as the stimulation level increases, as evidenced by the steeper slopes on the y-axis of Figure 8.

**Figure 8:**
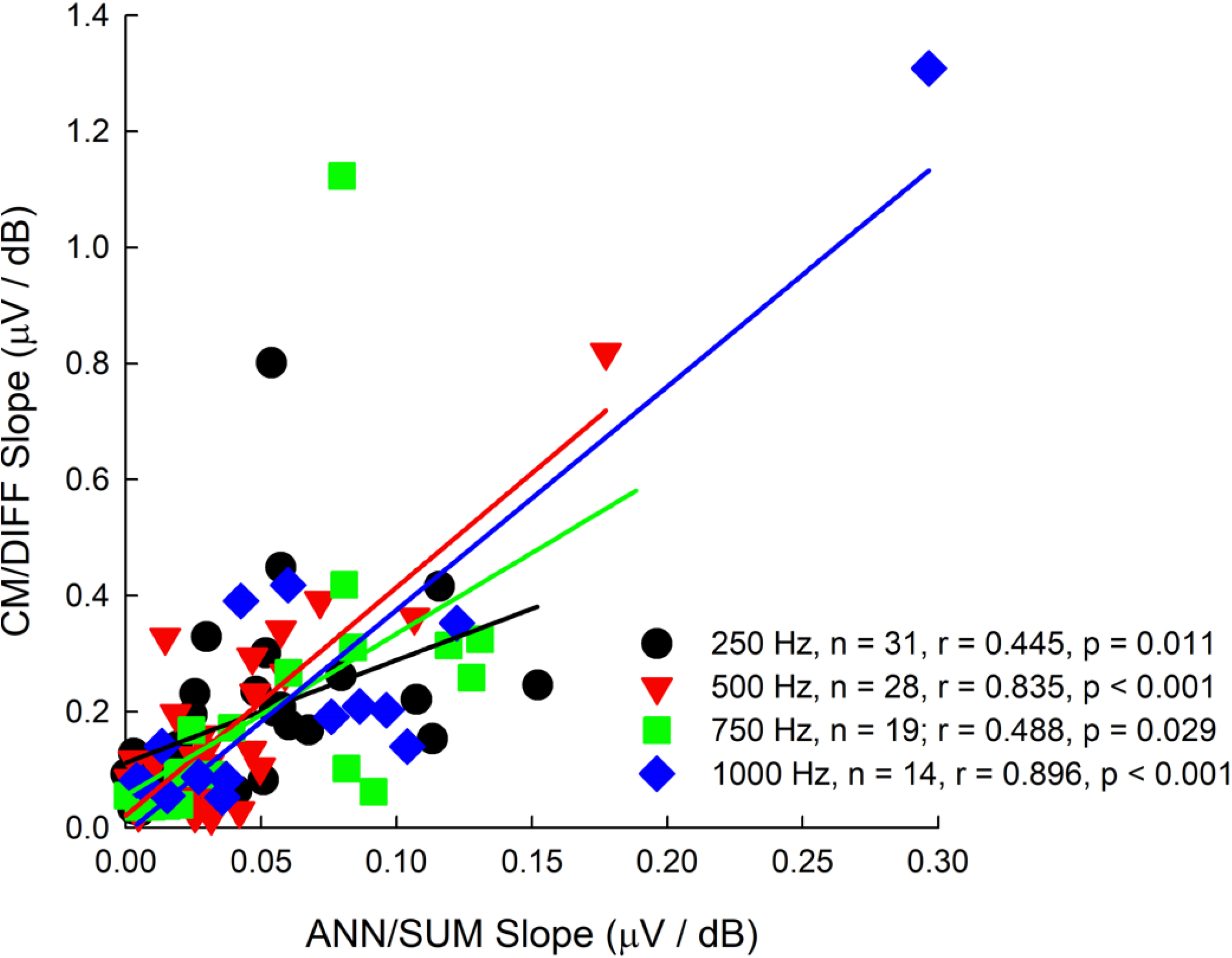
Correlations between CM/DIFF and ANN/SUM amplitude growth function slopes for 250, 500, 750, and 1000 Hz stimuli.

## DISCUSSION

Past studies of electrocochleography in CI users have focused on its application during CI surgery (Bester et al, 2022; Lenarz et al, 2022), to predict post-operative speech understanding and behavioral audiometric thresholds (Abbas et al, 2017; Koka et al, 2017, Fontenot et al, 2019), and to guide programming of the hearing aid component of the EAS sound processor (Agrawal et al, 2020). The current study provides a new application of ECoG, that is, to be used as an objective measure of longitudinal peripheral auditory function in EAS CI users. The stability of both CM and ANN amplitudes seen in our Nucleus CI users is consistent with the intertest and intratest reliability of ECoG measures in normal hearing and sensorineural hearing loss populations (Bergholtz et al, 1976; Densert et al, 1994; Mori et al, 1981; Park & Ferraro, 1999). Thus, the increase in ECoG thresholds and decrease in ECoG amplitudes in those with delayed hearing loss is likely clinically meaningful. The slope of the ECoG amplitude growth function was not a meaningful metric, as slopes were stable across repeated test sessions for both subjects with stable hearing and subjects with delayed loss of hearing.

One limitation of ECoG studies in general is that the difference / summation technique to separate out hair cell / neural potentials is not perfect. That is, it is very possible that there is still a neural component in the CM/DIFF traces that hasn’t been isolated out (e.g. Forgues et al 2014; Abbas et al, 2017). The contamination of the ANN/SUM responses by the CM can be seen at stimulus levels 30-40 dB above CM threshold (Forgues et al 2014). Thus, for subjects with particularly large dynamic ranges in their ECoG amplitude growth functions, some of the suprathreshold amplitudes may be affected by this contamination.

From a clinical perspective, both CM and ANN potentials are equally affected with loss of hearing, as shown by Figure 4 and Table 4. However, the threshold measures may be more a more reliable indicator of loss of hearing as they are not as affected by incomplete separation of CM and ANN potentials using the difference and summation techniques (Forgues et al, 2014). Regardless, the similar impact on CM and ANN thresholds and slopes implies a common underlying reason of delayed hearing loss, though there are many probable etiologies that are debatable. Eshraghi et al. (2013) observed outer hair cell loss in their animal models of CI, which at first glance appears consistent with loss of CM and ANN potentials in our delayed-hearing loss population. However, their animal CI protocol involved extensive electrode-induced cochlear trauma such that shifts in both low- and high-frequency hearing were observed. These conditions may not necessarily reflect our low-frequency hearing preservation patients.

Animal studies have shown that the ANN/SUM potential is sensitive to changes in endocochlear potentials (Lichtenhan et al, 2017). Compromised endocochlear potentials have been implicated in delayed hearing loss, at least in the high-frequency region of the cochlea where the CI electrode lies (Tanaka et al. 2014; Reiss et al. 2015). Alternatively, our previous human electrophysiological findings based on complex electrode impedance measures, as well as a post-mortem human EAS CI study, suggest intracochlear fibrosis as a contributor to delayed hearing loss (Quesnel et al, 2016; Tejani et al, 2022).

In general, animal histology has suggested many possible etiologies, such as compromised endocochlear potentials (Tanaka et al. 2014; Reiss et al. 2015), cochlear neuropathy/synaptopathy (Li et al. 2020), and excitotoxicity (Kopelovich et al. 2015). A common theme is that these studies in general do not implicate hair cell / neural damage / pre-synaptic / post-synaptic damage (O’Leary et al. 2013; Tanaka et al. 2014; Reiss et al. 2015; Quesnel et al. 2016). Without post-mortem histology, one cannot definitely prove the underlying causes of delayed hearing loss in humans post-CI (e.g. Quesnel et al, 2016). This limitation thus makes electrophysiology, including electrocochleography, a valuable tool to investigate loss of hearing in human CI users (e.g. Scheperle et, 2017; Tejani et al, 2021, 2022).

Our secondary analysis of correlations with audiometric thresholds shows that regardless of audiometric frequency, there is a strong correlation between ECoG thresholds and audiometric thresholds. This is despite the fact that a combination of electrode arrays was used, with some electrodes having partial insertions. These strong correlations are consistent with past studies conducted by our institution (Abbas et al, 2018; Kim et al, 2018) and others (Koka et al, 2017; Agarwal et al, 2021). In contrast, our slope metrics were generally not well correlated with behavioral audiometry. Additionally, subjects with greater residual hearing also tended to have higher ECoG amplitudes, which is consistent with correlations between ECoG “total-response” amplitudes and behavioral audiometry (Fitzpatrick et al, 2014). This was especially the case for CM/DIFF potentials. In the case of the ANN/SUM amplitudes, these correlations were not statistically significant in some cases. The lack of correlation may stem from the lower spread of ANN/SUM amplitudes since these potentials are generally small.

Our present study provides a new application of post-operative ECoG in monitoring residual hearing and understanding the pathophysiology of delayed hearing loss. In a future study, we will explore further applications of post-operative ECoG, including the relationship between post-operative ECoG and speech perception, as most data in the literature focuses interoperative ECoG at time of surgery and correlating that with outcome measures (Fontenot et al, 2019, Canfarotta et al, 2021; Walia et al, 2022). In addition, while our measures were conducted using in-house custom-built equipment, cochlear implant companies are also designing and implementing hardware and software adaptations so that their own systems can be used for ECoG recordings. Thus, it is foreseeable that both intraoperative and post-operative ECoG recordings can be integrated into clinical practice.

## Data Availability

All data produced in the present study are available upon reasonable request to the authors

